# Inter-laboratory comparison of HbA1c can effectively improve test consistency of primary medical institutions

**DOI:** 10.1101/2024.12.27.24319677

**Authors:** Pan Liping, Yang Jun, Luo Kun, Yun Hao, Wei Shen, Tan Jin, Lin Ming

**Affiliations:** Laboratory Department, Wuhan Center for Clinical Laboratory, Wuhan, Hubei, China; Clinical Laboratory, Wuhan Asia Heart Hospital, Wuhan, Hubei, China; Union Hospital, Tongji Medical College, Huazhong University of Science and Technology, Wuhan, Hubei, China

**Keywords:** inter-laboratory comparison, HbA1c test, consistency, primary medical institutions, mutual recognition of medical tests

## Abstract

**Background:** Glycated hemoglobin (HbA1c) represents a diagnostic index and an important long-term measure for monitoring Diabetes mellitus (DM). In this study, we investigated the quality of HBA1c test in primary medical institutions, and conducted a comparison project of HBA1c test for two years, analyzed and evaluated the test consistency of HBA1c.

**Methods:** A cohort study was conducted by Wuhan Center for Clinical Laboratory, involving 51 primary medical institutions and 29 tier-three hospitals. Fresh frozen blood samples were prepared by a clinical laboratory certificated by the National Glycohemoglobin Standardization Program (NGSP), and distributed to 80 participating institutions every three months. Microsoft Excel 2021 software was employed to analyze the mean, standard deviation and coefficient of variation of the test results.

**Results:** Results showed that 60.8% HBA1c test results cannot reach the quality requirements of HbA1c measurement (CV<3.5%) in primary medical institutions. After executing the project, the laboratory pass rate of HBA1c test increased from 39.2% to 64.6%, and the maximum coefficient of variation (CV) within the immunological assays of HBA1c test dropped from 18.0% to 11.4%.

**Conclusions:** Inter-laboratory comparison of HbA1c can effectively improve test consistency of primary medical institutions. The finding has implications for diabetes management strategies, emphasizing the importance of continuous comparison of HBA1c test in primary medical institutions, providing a practical example to accelerate the mutual recognition of medical test results in a wider range.

## 1. Introduction

Diabetes Mellitus (DM) is a condition that poses a heavy burden on public health systems and individuals across the globe. A World Health Organization (WHO) report stated that the mortality rate due to DM rose by 3% from 2000 to 2019 [1], and DM is the leading cause of 1.5 million deaths in 2019 [2]. Glycated hemoglobin HbA1c is formed by irreversible nonenzymatic glycation, reflects the average blood glucose over recent 2 to 3 months [3]. HbA1c is formally become a diagnostic measure (HbA1c 6.5%) for diabetes in 2011, and is considered a key parameter for monitoring the progression of diabetes [4]. However, more than 30 different HbA1c measurement devices based on various methods have been developed and applied in medical laboratories, sometimes it resulted in incomparable results [5, 6]. Consistent and comparable results from different laboratories are important for clinical practice guidelines to be applied to decisions about patient care. More importantly, as an indispensable part of routine management of diabetes, the quality and standardization of HbA1c test is key to enhancing the mutual recognition of test results among hospitals.

As the most important part of the hierarchical medical system, in many countries, primary medical institutions or primary care physicians have played a very important role in patient care and rehabilitation of most chronic diseases, including DM. The longitudinal use of the HbA1c test requires strict quality management including accreditation of the laboratory, a dedicated internal control design, participation in an external quality assessment (EQA) program (proficiency test), and careful consideration of pre- and post-analytical aspects of the test. However, according to the results of a previous study, the coefficient of variation (CV) of HBA1c test, which indicates the degree of variation of the test, between different laboratories (mostly tertiary medical hospitals) or different methods was as high as 20% or more [7]. Since, at present, no studies examined the quality of HBA1c test in poorly resourced primary medical institutions, presumably, the situation might be worse. Our research team carried out an inter-laboratory comparison program for HbA1c detection in Wuhan from 2022 to 2023 (January 2022-December 2023), and evaluated the consistency of the results of HBA1c detection among various laboratories, with an aim to provide a basis for improving the quality of DM diagnosis in primary medical institutions and promoting the clinical use of HBA1c.

## 2. Materials and Methods

### 2.1 Participants and procedures

Participating institutions included 80 laboratories under 51 primary medical institutions and 29 tertiary medical institutions in Wuhan. Participating laboratories received 10 free samples every three months, and reported the results to the External Quality evaluation (EQA) platform of Wuhan Center for Clinical Laboratory as scheduled.

### 2.2 Specimen preparation

Patient samples were remaining samples for clinical examination, collected by the laboratory of Asian Cardiology Hospital every three months, with the HbA1c concentration ranging from 5.0% to 10.0%. Abnormal hemoglobin (Hb) specimens were eliminated by hemoglobin electrophoresis to ensure that abnormal Hb < 5%. After preparation, they were stored at minus 80 °C . Samples within the same concentration range were mixed, 50 ml resultant specimens were prepared and then packaged at 100 μ l/specimen. Packaged samples, after quality checking against state standards, were then distributed to all participating laboratories.

This is a retrospective study of remaining samples for clinical examination. Patient samples were collected from Dec 1, 2021 to Sep 15, 2023, relevant data of HbA1c concentration is obtained after collected. All collected samples do not involve any patient information or medical information, only include data of HbA1c concentration; all authors had no access to information that could identify individual participants during or after data collection. Ethics Committee of Wuhan Center for Clinical Laboratory approved the study (No. WHCCL Ethics Committee ❲2023❳ 003). Verbal informed consent was obtained through telephone, one researcher documented and another one witnessed the obtaining process to make sure that all sample sourced participants confirmed the statement “I agree that my samples can be used for subsequent clinical research”.

### 2.3 Determination of target values

The target value of the specimens was determined by the laboratory of Asian Cardiology Hospital, a Level I Laboratory certificated by National Glycohemoglobin Standardization Program (NGSP), by using High-performance liquid chromatography (HPLC) for HbA1c measurement. The NGSP HbA1c standardization program began in 1996, and is committed to promoting the standardization and traceability of HbA1c testing worldwide. As an NGSP-certificated Level I laboratory, Asian Cardiology Hospital was monitored on a quarterly basis via the exchange of 10 fresh frozen samples and has been passing the NGSP criteria on each assessment session in order to maintain their certification status. The testing instrument used is Variant II Turbo of Bole (instrument No. YXLM-GEN-A.03), the testing reagents included Bole’s original reagent, calibration products kits and quality control sets. The instrument is regularly maintained and calibrated according to the laboratory’s standard operating procedure (SOP).

After assigning a value to the whole blood specimens, the specimens were distributed to participating laboratories in 24 hours via cold chain transportation at -20 °C . The preliminary experimental data showed that the uniformity and stability of the samples met the requirements under this transportation condition.

### 2.4 Consistency Comparison

Consistency comparison was made every quarter, and results were not grouped according to instruments or methods. Laboratories must submit testing data within a week after receipt of specimens on quarterly basis. The consistency criteria required that the deviation of the test result from the target value should be less than or equal to 6%. More than 80% of the results satisfying the criteria was considered acceptable. After quarterly comparison, field guidance was provided and instrument calibration was carried out for unqualified laboratories.

### 2.5 Data Collection

Data were uploaded to the EQA platform of Wuhan Clinical Laboratory Center.

### 2.6 Statistical Analysis

Data were analyzed using Microsoft® Excel® 2021MSO (version 2409 Build 16.0.18025.20030), including methodological distribution, pass rate, and variability. Single sample K-S test was utilized for normality test, and a P>0.05 was considered to be normal distribution. The normally-distributed data were expressed as z±s, and the paired t-test was used for inter-group comparisons. Chi-square test was used to compare the pass rate among different levels of medical institutions. A P<0.05 was considered to be statistically significant.

## 3. Results

### 3.1 Detection Methods Used by Institutions

Various methods were used for HbA1c determination, including Ion-Exchange High-Performance Liquid Chromatography (IE-HPLC), Boronate-Affinity HPLC, immunoassay, enzymatic assay, Point-of-care Testing (POCT), among others.

In tertiary medical institutions, HPLC was a primary technique for HbA1c detection. Of the 29 tertiary medical institutions, 23 (79.3%) employed HPLC-ion exchange, 3 institutions (10.7%) used HPLC-borate affinity, and 3 institutions utilized other methods, such as immunoturbidimetry or enzymatic assays.

In primary healthcare institutions, a variety of instruments and methodologies were used. Of the 51 primary healthcare institutions, 20 employed HPLC-ion exchange or borate affinity (39.2%), 19 institutions used immunofluorescence or immunoturbidimetry (37.3%), and 12 utilized liquid chromatography and POCT, as shown in figure 1.

**Figure 1.**
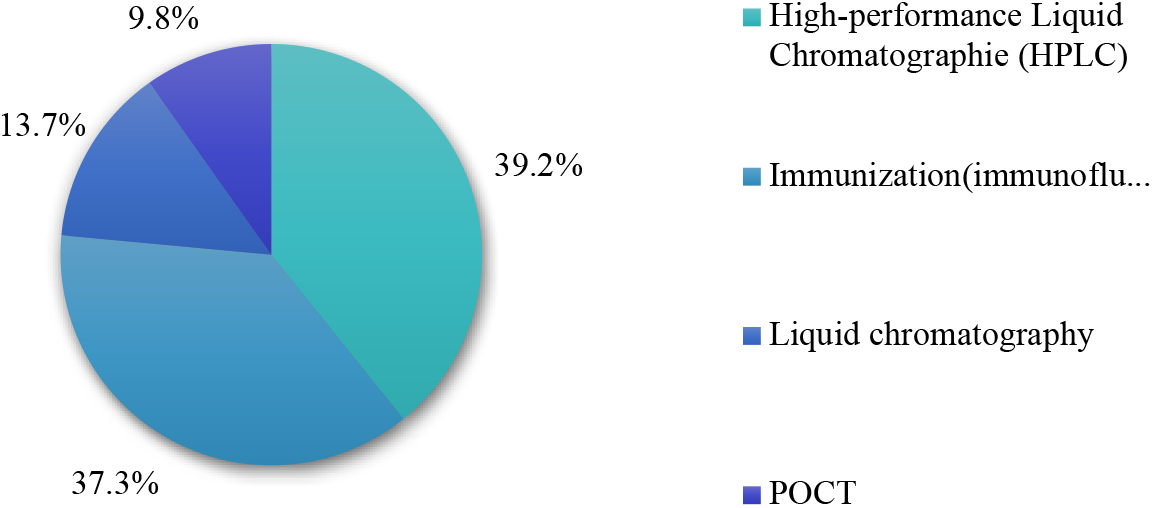
Methods used in primary healthcare institutions

### 3.2 Pass rate of laboratories

Against the passing criteria, the pass rate of tertiary medical institutions was significantly higher than that of primary medical institutions (P<0.05). In the first quarter of 2022, the pass rate of primary care institutions was only 39.2%, and by the fourth quarter of 2023, the pass rate increased to 64.6%, as shown in Figure 2.

**Figure 2.**
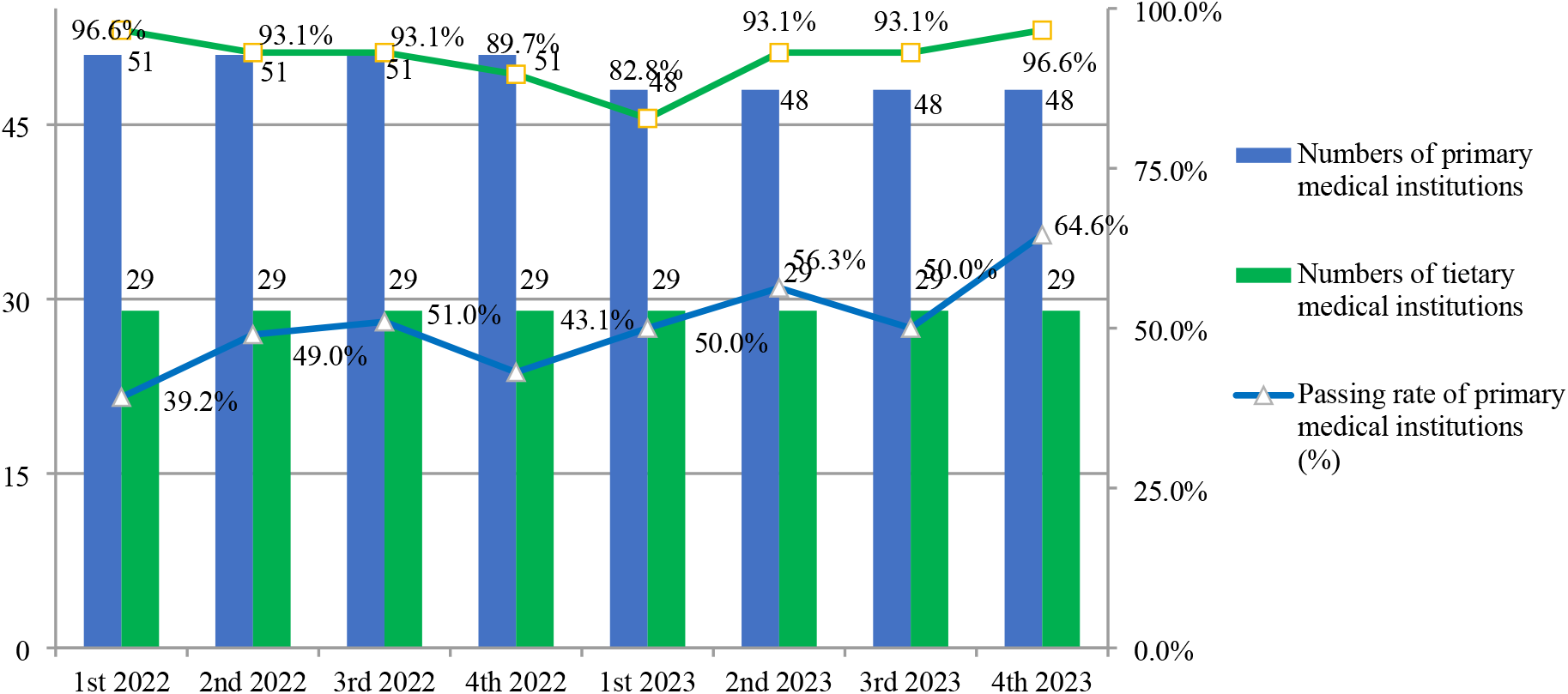
* Passing rate of different medical institutions in terms of HBA1c consistency * The horizontal axis represents chronological order of the comparison project. The left and right vertical axis displays the numbers of laboratories and passing rate of HBA1c test, respectively.

### 3.3 Differences in methods within groups

Results obtained by four different methods, involving 3 different representative concentrations (6.0-7.0%, 7.5-8.5%, 9.5-10.5%), in 51 primary medical institutions were analyzed. Among the four methods, the variation in POCT group was greatest, the coefficient of variation (CV) being up to 19.8%. From the first quarter of 2022 to the fourth quarter of 2023, the CVs of four methods group all decreased significantly, as exhibited in Table 1.

**Table 1.**
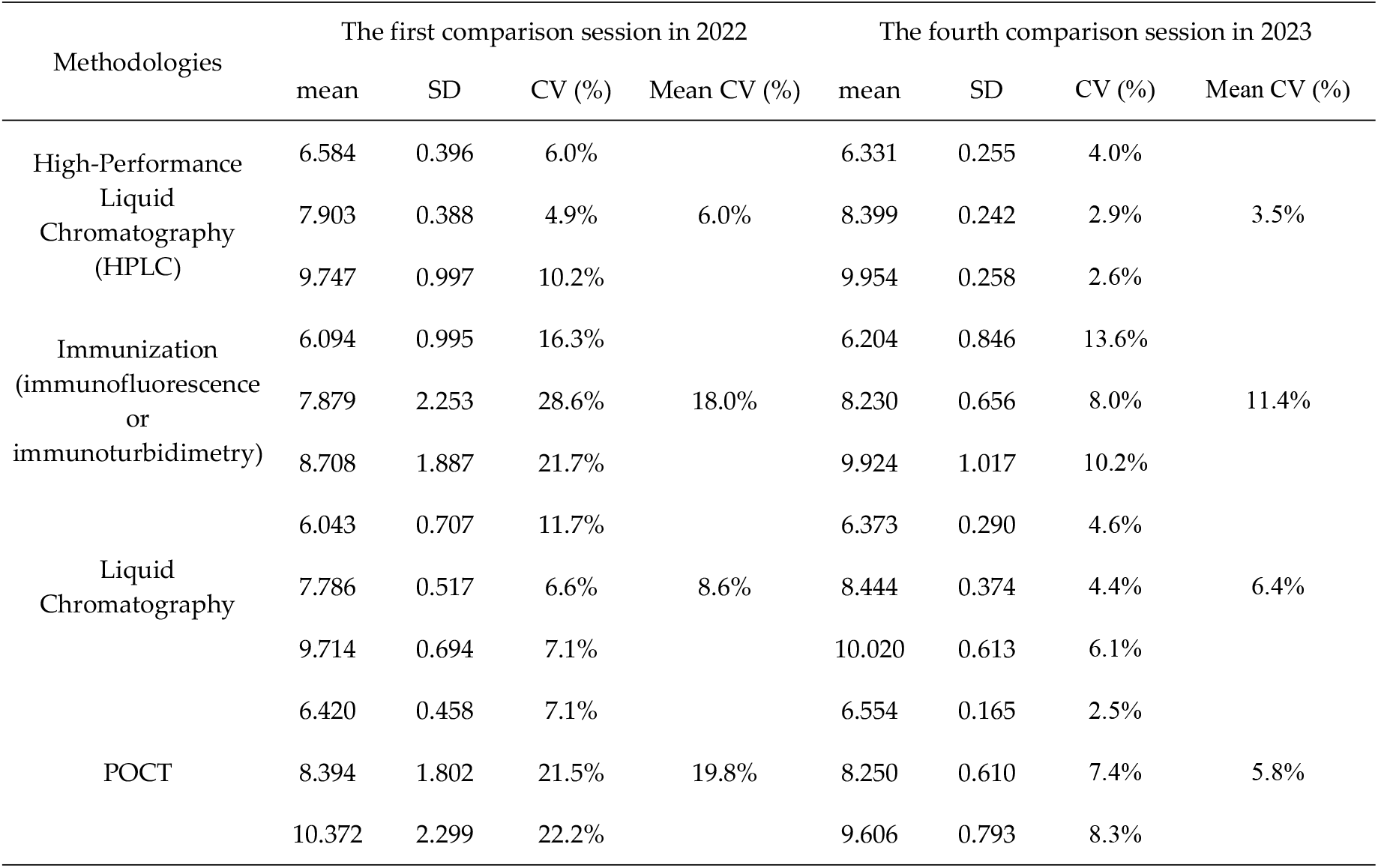
Bias and variation among different detection methods in primary medical institutions.

### 3.4 Coefficient of variation of immunoassay

In this study, 19 primary medical institutions used immunological methods for HBA1c detection, and the variability of inter-laboratory detection varied substantially. From the first quarter of 2022 to the fourth quarter of 2023, the variability of inter-laboratory detection gradually dropped, from 18.0% to 11.4%, as shown in Figure 3.

**Figure 3.**
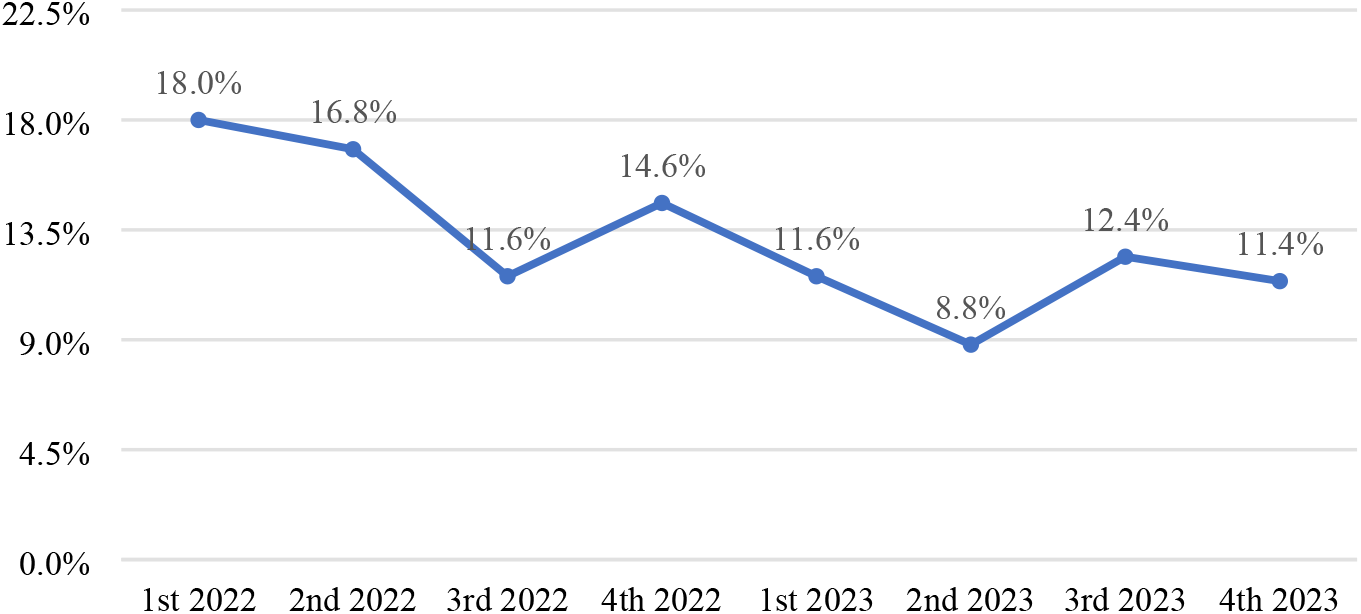
Coefficients of variation of immunoassays* * The horizontal axis represents chronological order of the comparison project, and the vertical axis displays the coefficients of variation.

## 4. Discussion

The clinical application of a testing metric is intimately related to its measurement quality or accuracy. The international HbA1c standardization program, starting in 1996, was designed to make the test results of the same analyte clinically consistent among different laboratories and different detection systems through specimen comparison [8]. In China, the standardization of HbA1c measurement commenced in 2020, a project known as “China Glycohemoglobin Standardization Program” (CGSP). CGSP originated from Shanghai Glycohemoglobin Standardization Program in 2010, setting up the qualification standard at deviation ≤ 6% of target value. In this study, we applied the CGSP consistency criteria (deviation ≤ 6%), and the overall pass rate (84.5%) of HBA1c measurement of tertiary medical institutions was significantly higher than that (50.2%) of primary medical institutions, multiple factors might have contributed to this finding. As we all known, only persistent and joint effort is needed to continuously improve the measurement quality of HbA1c and provide a more reliable bedrock for clinical management of DM.

Primary care serves as the cornerstone in a strong healthcare system. In recent years, primary healthcare institutions have undergone continuous reforms, striving to improve their efficiency and performance. However, they still grapple with financial constraints, a dearth of skilled professionals, sluggish departmental growth, and limited capacity to treat major diseases [9]. In our previous survey, of all 188 primary medical institutions in Wuhan, 60.0% had two or fewer laboratory personnel, 27.1% performed HbA1c test, and 60.8% test results cannot reach the quality requirements (CV<3.5%). Daily clinical duties make it hard for these primary medical institutions to send staff for advanced professional training. As a result, many laboratories (accounting for 53.4% in Wuhan) have failed to establish a quality management system. Many laboratory staff lack quality control awareness, are foreign to instrument operation, do not understand the working principles of HbA1c determination. With the implementation of the inter-laboratory comparison program, the research team provides specific guidance to unqualified laboratories on quarterly basis, and the pass rate has been improved (having risen from 39.2% to 64.6% in primary medical institutions).

In this study, a variety of instruments and methodologies were used in primary healthcare institutions, and results showed that, among the four methods, the variation coefficient of immunoassay and POCT group was really high, sometimes more than 20.0%. Previous study reported that some techniques and instruments were gradually phased out because of unstable HbA1c test results [8]. In recent years, with the upgrading of detection equipment, HPLC method is extensively used in tertiary medical institutions, while immunoturbidimetric essays and point-of-care test (POCT) are still very common in primary medical institutions. Apart from lack of professionals, the small amount of detection, high maintenance cost and expense of quality control products are important factors leading to this situation. HPLC is an automatic detection method with high reliability and accuracy, and has been seen as a state-recommended method for HbA1c detection [10]. On the contrary, immunoturbidimetric detection requires pre-treatment, which is greatly subject to operator skill and might result in undesirable reproducibility and accuracy in HbA1c measurement [11]. POCT is naturally a wise choice for primary medical institutions since it requires little expertise and easy to operate with no major procedural challenges [12]. However, according to Expert Consensus Committee on HbA1c measurement, the precision and accuracy of most POCT methods, at present, fail to meet clinical needs [13,14], and thus cannot be used for diabetes diagnosis [15], and are recommended as a monitoring test. Some studies reported that under strict and standardized laboratory quality control, including standard operating procedures, internal quality control (IQC), regular instrument calibration, etc. the performance will meet the criteria for clinical application in terms of precision and accuracy [16]. In this study, coefficient of variation (CV) within all four method groups was lowered at the end of the comparison program. For the immunological assay group, CV decreased from 18.0% to 11.4%. According to the quality requirements of HbA1c determination (CV<3.5%), HbA1c detected by HPLC in primary medical institutions can satisfy the clinical requirements at present, but it’s recommended that the results using other methods should only serve as an auxiliary indicator for diabetes management, and there is still a long way to go before primary clinical institutions can employ these methods with precision and accuracy that are up to the requirements of clinical application.

External quality assessment (EQA) is an effective way to verify the reliability and comparability of laboratory test results, and is also a measure indicative of the result of standardization work [10]. However, due to insufficient commutability, most processed EQA materials are unsuitable to assess trueness of HbA1c assays and agreement between different assays or laboratories [17]. The samples used in the evaluation should be as close as possible to clinical samples, which is very important for the screening of diabetes, the formulation of blood glucose management protocol and the evaluation of the efficacy. In this study, we used fresh whole blood samples with no obvious matrix effect as comparison specimen. The project improved the consistency of HbA1c tests in primary medical institution, provided an example for improving the detection performance and diabetes management, and offered a practical example to accelerate the mutual recognition of medical test results in a wider range.

This study is subject to several limitations. First, only one laboratory was chosen to determine the value of comparison specimens. In future, we will cooperate with more capable and qualified partners in promoting the consistency of HbA1c measurement in primary medical settings. In addition, HbA1c test is not widely used in diabetes management in primary medical institutions at present, only 27.1% institutions performed this item, and too many detection methods resulting in a small number in some method groups. As a stable monitoring index, HbA1c is recommended to be included in the diabetes management plan in primary medical settings under the premise of standard quality control.

## Conclusion

Inter-laboratory comparison of HbA1c can effectively improve test consistency of primary medical institutions. The finding has implications for diabetes management strategies, emphasizing the importance of continuous comparison of HBA1c test in primary medical institutions, providing a practical example to accelerate the mutual recognition of medical test results in a wider range.

## Data Availability

All relevant data are within the manuscript and its Supporting Information files.

## Acknowledgments

We are indebted to Asian Cardiology Hospital, all primary medical institutions and our research team members for participating in this study.

